# Serial measurements in COVID-19-induced acute respiratory disease to unravel heterogeneity of the disease course: design of the Maastricht Intensive Care COVID cohort; *MaastrICCht*

**DOI:** 10.1101/2020.04.27.20080309

**Authors:** Maastricht Intensive Care COVID Study Group; *MaastrICCht* Collaborators, Jeanette Tas, Rob J.J. van Gassel, Serge J.H. Heines, Mark M.G. Mulder, Nanon F.L. Heijnen, Melanie J. Acampo-de Jong, Julia L.M. Bels, Frank C. Bennis, Marcel Koelmann, Rald V.M. Groven, Moniek A. Donkers, Frank van Rosmalen, Ben J.M. Hermans, Steven J.R. Meex, Alma M.A. Mingels, Otto Bekers, Paul H.M. Savelkoul, Astrid M.L Oude Lashof, Joachim E. Wildberger, Maastricht Intensive Care COVID Study Group, Fabian H. Tijssen, Wolfgang F.F.A. Buhre, Jan-Willem E.M. Sels, Chahinda Ghossein-Doha, Rob G.H. Driessen, Pieter L. Kubben, Marcus L.F. Janssen, Gerry A.F. Nicolaes, Uli Strauch, Zafer Geyik, Thijs S.R. Delnoy, Kim H.M. Walraven, Coen D.A. Stehouwer, Jeanine A.M.C.F. Verbunt, Walther N.K.A van Mook, Susanne van Santen, Ronny M. Schnabel, Marcel J.H. Aries, Marcel C.G. van de Poll, Dennis C.J.J Bergmans, Iwan C.C. van der Horst, Sander M.J. van Kuijk, Bas C.T. van Bussel

## Abstract

**Background:** The course of the disease in severe acute respiratory syndrome coronavirus 2 (SARS-CoV-2) infection in mechanically ventilated patients is unknown. To unravel the clinical heterogeneity of the SARS-CoV-2 infection in these patients, we designed the prospective observational Maastricht Intensive Care COVID cohort; *MaastrICCht*. We incorporated serial measurements that harbour aetiological, diagnostic and predictive information. The study aims to investigate the heterogeneity of the natural course of critically ill patients with SARS-CoV-2 infection.

**Study population:** Mechanically ventilated patients admitted to the Intensive Care with SARS-CoV-2 infection.

**Main message:** We will collect clinical variables, vital parameters, laboratory variables, mechanical ventilator settings, chest electrical impedance tomography, electrocardiograms, echocardiography as well as other imaging modalities to assess heterogeneity of the natural course of SARS-CoV-2 infection in critically ill patients. The *MaastrICCht* cohort is, also designed to foster various other studies and registries and intends to create an open-source database for investigators. Therefore, a major part of the data collection is aligned with an existing national Intensive Care data registry and two international COVID-19 data collection initiatives. Additionally, we create a flexible design, so that additional measures can be added during the ongoing study based on new knowledge obtained from the rapidly growing body of evidence.

**Conclusion:** The spread of the COVID-19 pandemic requires the swift implementation of observational research to unravel heterogeneity of the natural course of the disease of SARS-CoV-2 infection in mechanically ventilated patients. Our design is expected to enhance aetiological, diagnostic and prognostic understanding of the disease. This paper describes the design of the *MaastrICCht* cohort.

**Strengths and limitations of this study:**

- Serial measurements that characterize the disease course of SARS-CoV-2 infection in mechanically ventilated patients
- Data collection and analysis according to a predefined protocol
- Flexible, evolving design enabling the study of multiple aspects of SARS-CoV-2 infection in mechanically ventilated patients
- Single centre, including only ICU patients

## INTRODUCTION

Severe acute respiratory syndrome coronavirus 2 (SARS-CoV-2) infection is highly heterogeneous in its presentation.[1-3] Approximately 40% of patients show no clinical signs, 40% have a mild illness, whereas around 20% require hospitalisation, of whom 5-10% develop a critical disease that requires mechanical ventilation.[4] The SARS-CoV-2 disease course in mechanically ventilated patients is unknown, while the COVID-19 pandemic, caused by SARS-CoV-2, stresses Intensive Care resources to maximum capacity in pandemic areas such as the Netherlands.[5] Compared to other regions in the world[6], we had time to plan, with the advantage to design a study that investigates heterogeneity of the disease course over time.

We hypothesize that a comprehensive characterization of the heterogeneity of the natural course of critically ill patients with SARS-CoV-2 will enhance our aetiologic, diagnostic and prognostic understanding of the disease, which ultimately helps to guide Intensive Care resources and patient care. Therefore, we initiated the Maastricht Intensive Care COVID cohort; *MaastrICCht*. We intend to collect a broad set of clinical variables and biomarkers serially over time that precede the outcome in mechanically ventilated patients infected with SARS-CoV-2. Unfamiliarity with SARS-CoV-2 infection and its disease course raises many aetiologic, diagnostic and prognostic questions in Intensive Care practice. For example, how does lung compliance develop over the course of the infection? How does multi-organ failure develop over the course of the disease for patients that survive vs those that do not? What are the cardiovascular complications that can be diagnosed early? Which patients develop thrombosis?[7] Is thrombotic risk driven by inflammation and affected by comorbidities, such as obesity, type 2 diabetes mellitus and the presence of cardiovascular disease?[3, 7] Does immobilisation by a neuromuscular blockade, also, play a role?[8] Serial data to investigate such topics are scare.[9, 10]

The COVID-19 pandemic required swift implementation observational research activities to unravel the clinical changes in the course of the disease that precede favourable or worse outcome in mechanically ventilated patients with a SARS-CoV-2 infection admitted to the Intensive Care. We intend to use serial measurements to investigate the aetiology, diagnostic and prognostic value of respiratory variables (i.e. ventilator settings, prone positioning, chest electric impedance tomography (EIT))[11-14], cardiovascular variables (laboratory variables, electrocardiograms, echocardiograms and chest computed tomography (CT) scans)[10], metabolic variables (kidney function, liver biochemistry and electrolytes) and thrombotic complications (laboratory variables, thrombotic events) amongst others as described below. We describe the design of the *MaastrICCht* cohort in detail.

## METHOD AND ANALYSIS

### Participants

This prospective cohort study is conducted in patients admitted to the Intensive Care of the Maastricht University Medical Center (Maastricht UMC+), a tertiary teaching hospital in the southern part of the Netherlands. Usually, our Intensive Care Unit (ICU) has 27 beds, divided over three subunits to which all types of critically ill patients are admitted. However, to provide care for patients during the COVID-19 pandemic, our ICU underwent a rapid stepwise upgrade to a maximum of 64 beds, consisting of six subunits covering 52 beds for COVID-19 patients and two subunits covering 12 beds for Intensive Care patients without COVID-19. Therefore, our subunits of the ICU are located in the department of Anaesthesiology as well. The local institutional review board (Medisch Ethische Toetsingscomissie (METc) 2020-1565/ 300523) of the Maastricht UMC+ approved the study, which will be performed based on the regulations of Helsinki.

We intend to include all patients admitted to one of our six COVID IC subunits. Patients are intubated and mechanically ventilated and have signs and symptoms of a viral infection and a PCR positive for SARS-CoV-2 and/or a chest CT scan scored positive based on a CORADS-score of 4-5 by a radiologist.[15] Patients can be admitted via our emergency department, via non-ICU wards and by transportation from other ICUs either for tertiary care referral or due to lack of bed availability in the regional hospitals.

### Registry and research questions

We developed a protocol for this prospective cohort that contains variables of interest, based on existing literature and based on variables of interest of study initiatives from other centres and study groups (please see below).[16, 17] Many cohort studies in patients with COVID-19 collect similar variables. We sought to collect all of these variables to be able to address a broad range of research questions. We established our list of variables to be retrieved (supplemental **Table 1**), which we intend to extend in the future. Furthermore, we developed a uniform protocol for patient charts during admission to allow for uniform collection of variables. Thus, we designed the cohort study to register a baseline set of clinical, biochemical and COVID-19 specific variables, and to collect all relevant outcome variables. Additionally, we will record clinical data each day during ICU admission for patients included in the cohort.

A major advantage of this approach is that all variables collected for this cohort are part of routine clinical care but are now collected uniformly and reported in a predefined format. This prospective cohort will serve as an open-source database for other investigators to submit requests for data. The cohort steering committee will consider all requests. To foster data sharing in the future, we chose to align a major part of data collection with an existing Intensive Care data registry and two international COVID-19 data collection initiatives, in a way that is in line with the Findable, Accessible, Interoperable, and Reusable (FAIR) data principle.[18] Our design thereby enables the contribution of data to other initiatives. A data-sharing agreement and plan, approved by the local institutional review board, will be necessary to share data of the *MaastrICCht cohort* with other data collection initiatives. Also, the cohort is designed in such a way that additional measures can be added during the study period based on new knowledge obtained from the rapidly growing body of evidence. Approval by the local institutional review board will be necessary to add additional measurements that are not part of routine clinical care.

The first primary aim is to investigate the course of COVID-19 respiratory disease during mechanical ventilation. For this, we will use the ventilator setting and chest EIT. Additionally, we intend to investigate the role of prone positioning in the course of the disease. The second primary aim will be to describe and investigate cardiovascular changes over time that determine incident thrombosis during mechanical ventilation of COVID-19 patients. Here, we will focus on cardiovascular biomarkers, such as biochemical markers of inflammation and coagulation, and markers of cardiac structure and function (i.e. electrocardiogram, echocardiography and chest CT scans). Third, we will investigate the development of multi organ failure and compare its disease course for patients that survive vs those that decease. Research questions are organized according to the type of research, e.g. aetiology, diagnostic and prognostic, and examples are shown in **Table 1**.[5]

**Table 1.** Research questions in COVID-19 induced respiratory disease in an Intensive Care population on mechanical ventilation.

| **Research question** | **Aetiology** |
| --- | --- |
| 1 | Is the development of dynamic compliance and chest EIT parameters more favorable in survivors as compared to deceased? |
| 2 | Is the development of multi-organ failure worse in deceased as compared to survivors? |
| 3 | Is an increase in d-dimer, fibrinogen, C-reactive protein and ferritin plasma concentrations over time greater in patients with incident thrombosis? |
| **Diagnosis** |  |
| 4 | Do ECG changes over time discriminate between incident thrombosis? |
| 5 | Does SOFA score changes discriminate between death or survival? |
| 6 | Does prone, versus supine, positioning determine a favourable improvement in PF ratio over time? |
| 7 | Does EIT, dynamic compliance, ARDS PEEP/FiO<sub>2</sub> or clinical opinion diagnose optimal PEEP best? |
| **Prognosis** |  |
| 8 | Can conventional risk scores predict outcome in COVID-19 patients? |
| 9 | How do newly developed risk scores perform in our cohort?[19-23] |
| 10 | Can chest EIT predict prone positioning? |
| 11 | Can changes in PF ratio during the first 72 hours predict prone positioning? |
| 12 | Are serially measured coagulation tests able to predict adverse thromboembolic and mortality events during the course of the disease? |
| 13 | Does acute kidney injury burden predict mortality events during the course of the disease?[24] |

### Data collection

The design and serial data collection enable aetiological, diagnostic and prognostic research, and we will first focus on the topics of respiratory and cardiovascular disease and multi-organ failure. The flexible cohort design will allow us investigation of additional topics based on our extensive set of baseline and serial measurements (supplemental **Table 1**). All variables and measurements are predefined in our study protocol. Inclusion and collection of variables are performed by medical research interns and PhD candidates not involved in patient care. Data collection for this cohort has started on March 25^th^, 2020 and will be continued without a predefined end-date.

For the baseline characteristics, we will collect data that is aligned with the Dutch National Intensive Care (NICE) data registry and two international COVID-19 data collection initiatives. First, we will collect the minimal dataset of the NICE registry (https://www.stichting-nice.nl/), which includes data from each ICU across the Netherlands. The minimal dataset is the core registration that includes demographic, admission and discharge data of Dutch Intensive Care patients.[25] Second, we will collect data on the COVID-19 case report forms (CRF) of the International Severe Acute Respiratory and Emerging Infection Consortium (ISARIC) and World Health Organisation (WHO).[26] This ISARIC/WHO tool queries symptoms of clinical infection, comorbidities, pathogen testing, therapy and outcome variables. Third, we will collect data fostering the Cardiac complicAtions in Patients with SARS Corona vIrus 2 regisTrY (CAPACITY), which is an extension of the COVID-19 CRF and of the WHO list that additionally queries cardiac history, diagnostics and occurrence of cardiovascular complications in COVID-19 patients.[27] Fourth, by collecting a vast number of additional variables (both from records and monitors) uniformly, we will be able to merge our cohort to address additional questions in collaboration with others.

### Serial chest electrical impedance tomography

In our department, chest EIT is performed by specialist ventilation practitioners and is available if considered indicated to support clinical practice in patients with respiratory failure and thus is not performed in each patient each day.[14] During the upscaling of COVID-19 ICU subunits, the team of ventilation practitioners was extended with trained medical research interns and technical physicians to cover serial chest EIT measurements in each of the six COVID ICU subunits. Data collection on chest EIT was supervised by the specialist ventilation practitioners.

For the *MaastrICCht* cohort, our specialist ventilation practitioner team will start the first chest EIT measurement in each admitted patient after intubation as soon as clinically and logistically possible. We intend to obtain measurements in every mechanically ventilated COVID-19 patient with intervals of 2-3 days during their ICU admission, if feasible, and, in particular, after changing clinical conditions, i.e. a change in ventilator settings [28], clinical respiratory deterioration and changes in positioning (i.e. prone-supine and vice versa). If the patient stabilizes and receives pressure support ventilation, chest EIT measurement frequency will decrease, and one to two measurements will be done during the further course of the disease, while on mechanical ventilation. Our chest EIT measurement protocol has been aligned with the chest EIT protocol of the Erasmus University Medical Center, Rotterdam, The Netherlands, to enable pooling of chest EIT data in collaboration [11-14, 29], and is described in the **supplement**.

### Serial clinical, physiological and laboratory measurements

We intend to collect daily clinical and physiological variables, laboratory variables, electrocardiography, and medication use (please see supplemental **Table 1**).[30, 31] Consequently, a standard set of plasma and serum biomarkers will be measured daily. Also, for each serum blood sample, the leftover serum will be collected by our Clinical Chemistry department and stored for future biomarker studies.

### Additional measurements

We intend to collect data from echocardiography, chest X-rays and chest CT-scans performed in daily care.[32] The latter will be performed for primary pulmonary assessment (acquired as non-contrast triage scans) [33], or for dedicated cardiovascular assessment (based on contrast-enhanced CT scans).

### Outcome variables

We collect information on events and complications in all patients. We record variables on intubation and extubation, need for extracorporeal membrane oxygenation [34] and need for renal replacement therapy, thrombo-embolic events and resuscitation.[35] We collect data on time to discharge and need for readmission. In case a patient decease, an intensivist will define the cause of death.[36, 37]

### Data management

A customized electronic case report form (e-CRF) was developed and implemented for the current *MaastriCCht* cohort, using CASTOR (version 2020.18). Independent study monitoring of the organisation and the conduct of the study was in adherence to the Good Clinical Practice guidelines. All variables were considered in daily care and led to decisions on diagnostics and interventions.

### Outline of the statistical analyses plan

All reports will be written following reporting guidelines appropriate for the type of study (i.e., STROBE for observational studies, STARD for diagnostic studies, TRIPOD for prognostic studies; see the Enhancing the QUAlity and Transparency Of health Research network at www.equator-network.org for detailed information). We will describe baseline characteristics of the study sample as means ± standard deviation (SD), median and interquartile range (IQR), and percentages, as appropriate. P-values will be interpreted according to the American Statistical Association’s Statement on Significance and P-Values.[38] All external researchers submitting a data request will be required to produce a statistical analysis plan that will be reviewed by the cohort steering committee. A summary of the statistical analysis plan for the currently planned research questions is summarized below.

In case of incomplete variables, data will be imputed if the proportion of incomplete patients is over 5%, excluding longitudinal measures that will be analysed using generalized linear mixed-effects regression. Multiple imputation will be used with the percentage of incomplete patients as the number of imputations. Predictive mean matching will be used to draw values to be imputed, as this is more robust to misspecification of the imputation model. If applicable, the results of multiply-imputed data will be pooled using Rubin’s Rules. In other cases, results will be pooled using available pooling methods.

### Aetiological research questions

In order to use all serial data, we will use flexible longitudinal data analyses techniques, such as generalized linear mixed-effects models. Where applicable, we will model longitudinal and time-to-event data simultaneously using a joint model in case of survival endpoints compared to binary endpoints. First, we will categorize the sample into groups of predefined outcomes, such as survivor vs deceased, responder/non-responder to prone positioning; and presence vs absence of thrombotic events. Then, we will model the preceding serial data over time for these categories. After reporting the crude results of the generalized linear mixed-effects models, the models will be extended with covariates to adjust for potential confounders. Confounders will be retained in the model according to the method of Rothman[39], if they significantly contribute to the model, as quantified by the Akaike Information Criterion, or if they improve the precision of the estimated treatment effect. Effect-modification will be examined for sex and based on pre-specified hypotheses. Results are reported as effect size with 95% confidence intervals (CI).

### Diagnostic and prognostic research questions

Diagnostic and prognostic modelling will be performed with a minimum of ten events per variable that is regarded as a candidate diagnostic or prognostic variable. Variable selection will be performed using the Akaike Information Criterion to arrive at a more parsimonious model. Internal validation will be estimated using bootstrap resampling. In each bootstrap sample, the model fitting steps will be repeated. The bootstrap resampling yields a shrinkage factor that will be used to shrink the coefficients towards 0 to compensate for overfitting (i.e., the phenomenon that a model performs best on the data used to develop it). Also, the bootstrap internal validation will yield measures of performance adjusted for optimism (i.e., it estimates measures of performance in future patients). Model performance will be quantified using estimates of model fit (i.e. Nagelkerke’s R-squared), of discriminative ability (the area under the receiver operating characteristic curve, or AUC, including 95% CI), and calibration, by visually inspecting a calibration plot. If applicable, the model outcome will be dichotomized and test characteristics, such as sensitivity, specificity, positive and negative likelihood ratio, will be computed.

No previous studies exist that included combinations of serial measurements of clinical variables and biomarkers with outcome in mechanically ventilated COVID-19 patients in the ICU. The lack of previous literature makes it challenging to calculate sample size. In a pragmatic approach, with a world-wide requirement of understanding COVID-19 pathophysiology in mechanically ventilated patients, we chose a stepwise release of cohort data and locked the data collection for the first time at April 19^th^, 2020, to release the first data as timely as possible. The results of this first data collection will be reported shortly.

## FUTURE PERSPECTIVES

Data from this cohort will be released in a stepwise approach, so that in this acute phase most pressing research questions can be addressed, but that the accumulation of data will produce ever-larger datasets for future research projects. If the same hypothesis will be tested over multiple data exports, we plan to correct for sequential testing.[40] We also aim to analyse the electrocardiograms, echocardiography and CT imaging results that are acquired now for later analyses. In addition, the stored serum samples will be used for further biomarker research. We will collect data on extracorporeal membrane oxygenation, which can support the extracorporeal life support organization registry.[41] Also, we intend to analyse microbiology results as, for example, fast replication of RNA viruses is known to introduce mutations in viral replication. Replication mutations may introduce more or less virulent strains that infect humans. It remains unclear whether the most virulent SARS-CoV-2 strains of a total heterogeneous SARS-CoV-2 pool, in particular, are relevant for the course of the disease in infected patients who develop a critical illness.

Furthermore, the rapid spread of COVID-19 in the Netherlands required swift transfers of mechanically ventilated patients from smaller hospitals to our tertiary teaching hospital and from our hospital to Germany in several cases.[42, 43] The study design includes potential to address whether transferred patients’ disease course and outcome differ from other admitted patients. Moreover, regional differences in outcome, both within our national as in comparison to Belgium and Germany can be explored.

In addition, we plan a long-term follow-up via outpatient clinic visits to evaluate the quality of life of survivors and will collaborate with rehabilitation medicine specialists.[44] Later, when the COVID-19 pandemic recedes, we will modify the ongoing cohort data collection to investigate and observe pathophysiological changes in other clinical variables and novel biomarkers of interest that precede favourable and worse outcome in non-COVID-19 patients at the Intensive Care.

### Strengths and limitations

Our cohort study design has several strengths. First, the study is prospective by design and allows for many serial measurements in SARS-CoV-2 infected patients over time. Second, systematic data collection is performed using a predefined protocol. Next, the data collection is in line with the FAIR data principle and combines a national Intensive Care data registry and two COVID-19 data collection initiatives. Another strength is the flexible design of the cohort and the organisation. This allows extending the variable list soon. Later, the on-going cohort study’s focus can easily be redefined to allow investigation of new pathophysiological concepts using new techniques or biomarkers in relation to outcome of Intensive Care patients. Our study is a cross-departmental joint approach throughout Maastricht UMC+ and in the region. A limitation of the study is the single centre approach and thereby a relatively small sample size, although the sample size of certain research questions is enlarged by merging our data with data from other research groups, as is anticipated. Furthermore, the growing body of evidence during the COVID-19 pandemic might amend therapy over time and this could affect studying the course of the disease. However, on the patient level, we intend to collect data on therapy daily and this will shed light on the course of therapeutic alterations within the cohort over time. Also, the stepwise data release might impair the power of the initial analyses. However, the fast spread of the SARS-CoV-2 virus affects patients world-wide and requires data to guide clinical decisions.[6] Another limitation is that we include only patients admitted to the ICU.[45] Observations made in our study may be generalized to critically ill patients, only. To shed some light on possible selection mechanisms for ICU admission, we intend to compare our cohort with hospitalized non-ICU patients with SARS-CoV-2 infection. However, no further exclusion criteria are determined. Hence, a heterogeneous sample of patients admitted to the ICU is expected. Our inclusion strategy thereby reduces the chance of selection bias, which contributes to the internal validity of the result for mechanically ventilated COVID- 19 patients.

### Collaborations

We have described the study design and data collection. Our approach aims to combine and compare the cohort data with the Dutch NICE registry, with other COVID-19 data collection initiatives such as the ISARIC and WHO data collections, and with COVID-19 chest EIT data from the Intensive Care of the Erasmus University Medical Center, Rotterdam, the Netherlands among others.

## CONCLUSION

The COVID-19 pandemic requires the swift implementation of observational research activities to unravel the heterogeneity of the natural course of critically ill patients with SARS-CoV-2. Prospective serial uniform measurements will enhance our aetiologic, diagnostic and prognostic understanding of the disease and ultimately helps to guide Intensive Care resources and patient care. We have described the design and statistical analyses plan of the *MaastrICCht* cohort and plan to present its first results soon.

## Data Availability

Data availability is not applicable for the protocol.

## Authors’ contributions

Tas, vGassel, vdHorst, vKuijk, vBussel conceived and designed the study, and drafted the manuscript

Heines, Mulder, Acampo-de Jong, Bels, Bennis, Koelmann, Groven, Donkers, vRosmalen, Hermans, Meex, Mingels, Driessen, Kubben, Janssen, Delnoy, Schnabel were involved in the data collection process.

Heijnen, Bekers, Savelkoul, Oude Lashof, Wildberger, Tijssen, Buhre, Sels, Ghossein Doha, Nicolaes, Strauch, Geyik, Walraven, Stehouwer, Verbunt, vMook, vSanten, Aries, vdPoll, Bergmans, critically reviewed the manuscript.

All authors read and approved the final manuscript.

